# Humoral and cellular immune response against SARS-CoV-2 variants following heterologous and homologous ChAdOx1 nCoV-19/BNT162b2 vaccination

**DOI:** 10.1101/2021.06.01.21258172

**Authors:** Joana Barros-Martins, Swantje I. Hammerschmidt, Anne Cossmann, Ivan Odak, Metodi V. Stankov, Gema Morillas Ramos, Alexandra Dopfer-Jablonka, Annika Heidemann, Christiane Ritter, Michaela Friedrichsen, Christian Schultze-Florey, Inga Ravens, Stefanie Willenzon, Anja Bubke, Jasmin Ristenpart, Anika Janssen, George Ssebyatika, Günter Bernhardt, Jan Münch, Markus Hoffmann, Stefan Pöhlmann, Thomas Krey, Berislav Bošnjak, Reinhold Förster, Georg M. N. Behrens

## Abstract

Cerebral venous thrombosis was reported as a rare but serious adverse event in young and middle-aged vaccinees following immunization with AstraZeneca’s ChAdOx1-nCov-19 vaccine. As a consequence, several European governments recommended using this vaccine only in individuals older than 60 years leaving millions of ChAd primed individuals with the decision to either receive a second shot of ChAd or a heterologous boost with mRNA-based vaccines. However, such combinations have not been tested so far. We used Hannover Medical School’s COVID-19 Contact (CoCo) Study cohort of health care professionals (HCP) to monitor ChAd primed immune responses before and three weeks after booster with ChAd or BioNTech/Pfizer’s BNT162b2. Whilst both vaccines boosted prime-induced immunity, BNT induced significantly higher frequencies of Spike-specific CD4 and CD8 T cells and, in particular, high titers of neutralizing antibodies against the B.1.1.7, B.1.351 and the P.1 variants of concern of severe acute respiratory syndrome coronavirus type 2 (SARS-CoV-2).

## Main text

Tremendous worldwide efforts since the outbreak of the pandemic resulted in effective vaccines against SARS-CoV-2. The first European Medicines Agency (EMA) approved vaccine reaching market was the lipid nanoparticle-formulated mRNA vaccine BNT162b2 (Comirnaty; BNT) developed by BioNTech/Pfizer. BNT, encoding for the full length of SARS-CoV-2 structural surface glycoprotein (spike; S protein), was proven safe and 95% effective in preventing coronavirus disease 2019 (COVID-19) ^1^. Oxford University in collaboration with AstraZeneca developed ChAdOx1-nCov-19 (Vaxzevria, ChAd), a replication-deficient chimpanzee adenovirus-vectored vaccine encoding also the full-length of SARS-CoV-2 S protein. In the initial clinical trials, ChAd had an acceptable safety profile, albeit somewhat lower efficacy of 70.4% against symptomatic COVID-19 ^2^. These data, together with the efficacy of other vaccines including those from Moderna ^3^ and Johnson&Johnson ^4^, raised hopes for expeditious ending of the SARS-CoV-2 pandemic.

However, in the first half of March 2021, vaccinations with ChAd were abruptly halted due to increasing numbers of moderate-to-severe thrombocytopenia and unusual thrombosis cases, particularly cerebral venous thrombosis and splanchnic-vein thrombosis among vaccinees ^5^. This new syndrome, termed vaccine-induced thrombotic thrombocytopenia (VITT) ^6^, developed within 28 days after vaccination and was confirmed by a large population study in Denmark and Norway ^7^. Although exact mechanisms are still unclear, VITT appears to be induced by antibodies directed against platelet factor 4 that lead to platelet activation ^8^. Despite concerns, European Medicine Agency (EMA) concluded that ChAd vaccination benefits outweigh the potential risks for an individual (https://www.ema.europa.eu/en/news/astrazenecas-covid-19-vaccine-benefits-risks-context, accessed May 31^st^, 2021) and ChAd remains a valuable asset against COVID-19. However, many countries offered to vaccinees, who received the first ChAd dose, to choose between ChAd or mRNA-based vaccines as a second (boost) dose, despite lack of data showing safety, reactogenicity or immunogenicity of such heterologous prime-boost schedules ^9^.

Furthermore, mutations in SARS-CoV-2 caused the emergence of rapidly expanding variants, especially B.1.1.7 (British), P.1 (formerly named B.1.1.28.1; Brazilian), and B.1.351 (South African) variants ^10^, which raised concerns on the feasibility of containing the pandemic through vaccination. Antibodies induced by BNT and ChAd vaccines efficiently neutralize the B.1.1.7 variant, while the neutralization P.1 and B.1.351 variants seems to be reduced ^11–13^. Moreover, BNT vaccination has been shown to be about 13% and 28% less protective against development of symptomatic COVID-19 for variants B.1.1.7 and B.1.351, respectively ^14^. Similarly, it has been reported that protection from symptomatic COVID-19 following ChAd vaccination is slightly reduced for B.1.1.7 variant ^15^, while no protection against mild-to-moderate COVID-19 caused by B.1.351 variant was observed ^16^. It remains to be determined whether heterologous prime-boost regimens could induce equal or even stronger immune responses against the novel viral variants as compared to the homologous prime-boost regiments.

To analyze the efficacy of the heterologous prime-boost vaccination schedule, we used our CoCo cohort of HCP^17,18^ and monitored response to homologous and heterologous prime-boost COVID-19 vaccine treatment schedules. Vaccinees who received one dose of ChAd were, according to the current vaccination strategy in Germany, offered to choose between ChAd and BNT vaccines for a second dose. To determine immunogenicity of the homologous and heterologous immune regimens, we studied n=129 ChAd-primed vaccinees without previous SARS-CoV-2 infection, of which n=32 chose homologous and n=55 heterologous boosting. For comparison, we included a group of BNT/BNT vaccinated HCP. The vaccination and blood collection schedule is depicted in Fig. 1A with additional information (age, sex) in Extended Data Fig. 1A-C. A retrospective analysis revealed that anti-SARS-CoV-2 S IgG (anti-S IgG) and IgA had declined by 43% and 65%, respectively, from mean 30 days after ChAd prime to shortly before boosting, similar to declines in BNT/BNT vaccinated individuals (Extended Data Tab. 1A, B). Importantly, we found comparable levels of anti-S IgG and IgA antibodies against the S-protein in the ChAd/ChAd and the ChAd/BNT groups before booster indicating that both groups responded equally well after priming with ChAd.

**Fig. 1.**
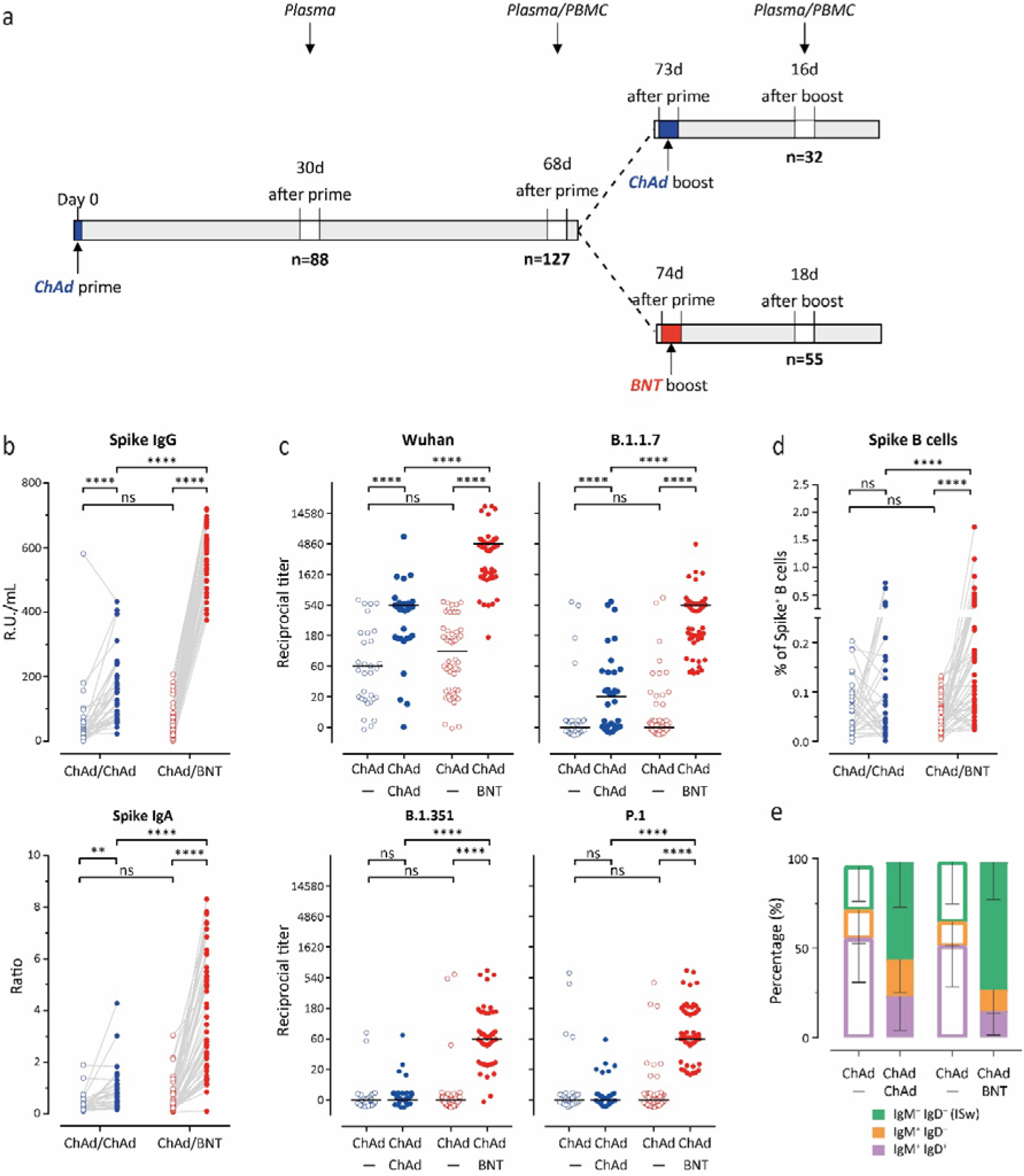
Stronger humoral immune response against all SARS-CoV-2 variants following heterologous ChAdOx1 nCoV-19 (ChAd) / BNT162b2 (BNT) than homologous ChAd / ChAd vaccination. a. Participant recruitment scheme. b. Spike (S)-specific IgG and IgA levels in plasma after prime (open circles) and after boost (closed circles) from homologous ChAd/ChAd (blue symbols) and heterolgous ChAd/BNT (red symbols) vaccinees. c. Reciprocal titers of neutralizing antibodies against Wuhan, B.1.1.7 (British), P.1 (B.1.1.28.1; Brazilian), and B.1.351 (South African) SARS-CoV-2-S variants measured using surrogate virus neutralization test (sVNT). d. Percentage of Spike-specific from total B cells in the whole blood measured using flow cytometry. e. Spike-B cells show isotype-switched (IgM^-^IgD^-^) phenotype following second immunization. Statistics: b. and d. **** p<0.001 Paired t test (within groups) or 2-way ANOVA followed by Sidak’s multiple comparison test (between groups); c. **** p<0.001 Chi-square test for trend.

Following the booster immunization, increased anti-S IgG and IgA responses were found in both groups. Heterologous ChAd/BNT vaccination led to a highly significant 11.5-fold increase for anti-S IgG as compared to a 2.9-fold increase after homologous ChAd vaccination (Fig. 1B, Extended Data Tab. 1C). We observed similar changes for anti-S IgA (Fig. 1B) indicating better humoral immune responses after heterologous prime/boost immunization. Anti-S IgG and IgA concentration after ChAd/BNT vaccination were within the range of fully BNT/BNT vaccinated individuals (Extended Data Tab. 1B and Extended Data Fig. 2A-B).

To test for neutralizing activity of antibodies induced by infection or vaccination, we recently developed an ELISA-based surrogate virus neutralization test (sVNT)^19^. In this assay, the soluble receptor for SARS-CoV-2, ACE2, is bound to 96-well-plates to which a purified tagged receptor binding domain (RBD) of the S-protein from the Wuhan strain (hCoV-19/Wuhan/Hu-1/2019) can bind once added to the assay. Binding is further revealed by an anti-tag peroxidase-labelled antibody and colorimetric quantification. Pre-incubation of the S-protein with serum or plasma of convalescent patients or vaccinees prevents subsequent binding to ACE2 to various degrees, depending on the amount of neutralizing antibodies present. Since we were interested to not only determine the neutralizing capacity of vaccination-induced antibodies against the Wuhan strain but also against some of the recently emerged variants of concern (VoC), we adapted the sVNT also to S proteins of the B.1.1.7, P.1, and B.1.351 variants. To validate these new assays, we applied sera from vaccinees that had been recently tested for their neutralizing capacity applying vesicular stomatitis virus (VSV)-based pseudotyped virus neutralization assays (pVNT)^12^. Comparing results obtained using pVNT with those of the newly developed sVNTs, we observed a high degree of correlation between both assays with R square values ranging between 0.50 and 0.69 (Extended Data Fig. 3). These findings demonstrate that the sVNT is suited to quantitatively assess the neutralization capacity of vaccination-induced antibodies not only against the Wuhan but also against the B.1.1.7, P.1, and B.1.351 variants of SARS-CoV-2.

Applying sVNT assays, we found that 81/88 participants possessed neutralizing antibodies against the Wuhan strain in pre-boost plasma. In contrast, neutralizing antibodies against the B.1.1.7 (17/88), P.1 (12/88), and B.1.351 (5/88) variants were less frequent (Fig. 1C; Extended Data Fig. 4). At 2-3 weeks after the booster immunization, frequencies and titers of neutralizing antibodies against the Wuhan strain increased in the ChAd/ChAd and the ChAd/BNT group with titers reaching higher values in the latter group (Fig. 1C; Extended Data Fig. 4). Differences between the ChAd and the BNT booster vaccination became even more evident when analyzing the neutralization capacity of antibodies induced against the VoC. In the ChAd/ChAd group booster immunization increased neutralization of the B.1.1.7 variant in some individuals but showed no effect against variants P.1 and B.1.351 (Fig. 1C; Extended Data Fig. 4). In contrast, booster immunization with BNT induced neutralizing antibodies at high frequencies against all analyzed VoC. In the ChAd/BNT group, all participants had neutralizing antibodies against the B.1.1.7 and P.1 variant and all but two participants possessed neutralizing antibodies against the B.1.351 variant (Fig. 1C; Extended Data Fig. 4). In the ChAd/BNT group the post boost neutralization capacity was highest against the Wuhan strain followed by the B.1.1.7 variant and less efficient against the P.1 and B.1.351 variant (Fig. 1C; Extended Data Fig. 4). Altogether, these data indicate that the booster immunization led to an increase of neutralizing antibodies in both vaccination groups and that the heterologous BNT booster vaccination efficiently induced neutralizing antibodies against all tested VoC.

We next determined the frequency and phenotype of B cells carrying membrane bound immunoglobulins specific for the S protein. PBMC were stained with 15 mAb, a viability dye, and an S-protein fused to a neo-green fluorescent protein (S-neoGreen; Extended Data Tab. 2) and analyzed by spectral flow cytometry (Ext. Data Fig. 5). Up to 0.2% of blood B cells in samples taken before booster vaccination were specific for the S-protein, with no significant difference between the ChAd/ChAd and the ChAd/BNT group (Fig. 1D open circles). In the ChAd/ChAd group, blood samples taken 2 to 3 weeks after the booster immunization did not reveal differences regarding frequencies of S-specific B cells compared to the pre-boost samples (Fig. 1D, filled dots). In contrast, S-specific B cells were strongly increased in the ChAd/BNT group following booster vaccination (Fig. 1D, filled dots). Furthermore, analysis of the IgM/IgD phenotype of the S-specific B cells revealed an increase in recently isotype switched B cells (IgD^-^IgM^-^) after booster immunization in both groups with higher frequencies in the BNT boosted group (Fig. 1E). The increased frequencies of isotype switched S-specific B cells after booster immunization went along with increased amounts of S-specific antibodies as well as increased neutralization capacities observed in both groups. Finally, the significantly increased overall frequency of S-specific B cells after BNT booster was paralleled by profound neutralization capacities against the VoC.

In addition to B cell-mediated immune responses, we also analyzed frequencies and phenotypes of S-specific T cells. To that end, density gradient-purified PBMCs were stimulated over night with DMSO alone or with pools of overlapping peptides dissolved in DMSO either covering the entire S-protein or the membrane (M), nucleocapsid (N), and the envelope (E) proteins. Cells were then stained with antibodies against cell surface molecules, fixed, permeabilized and then stained with antibodies against intracellular interferon (IFN)-γ and tumor necrosis factor (TNF)-α Frequencies of (IFN)-γ and (TNF)-α were determined by flow cytometry (Fig. 2A).

**Fig. 2.**
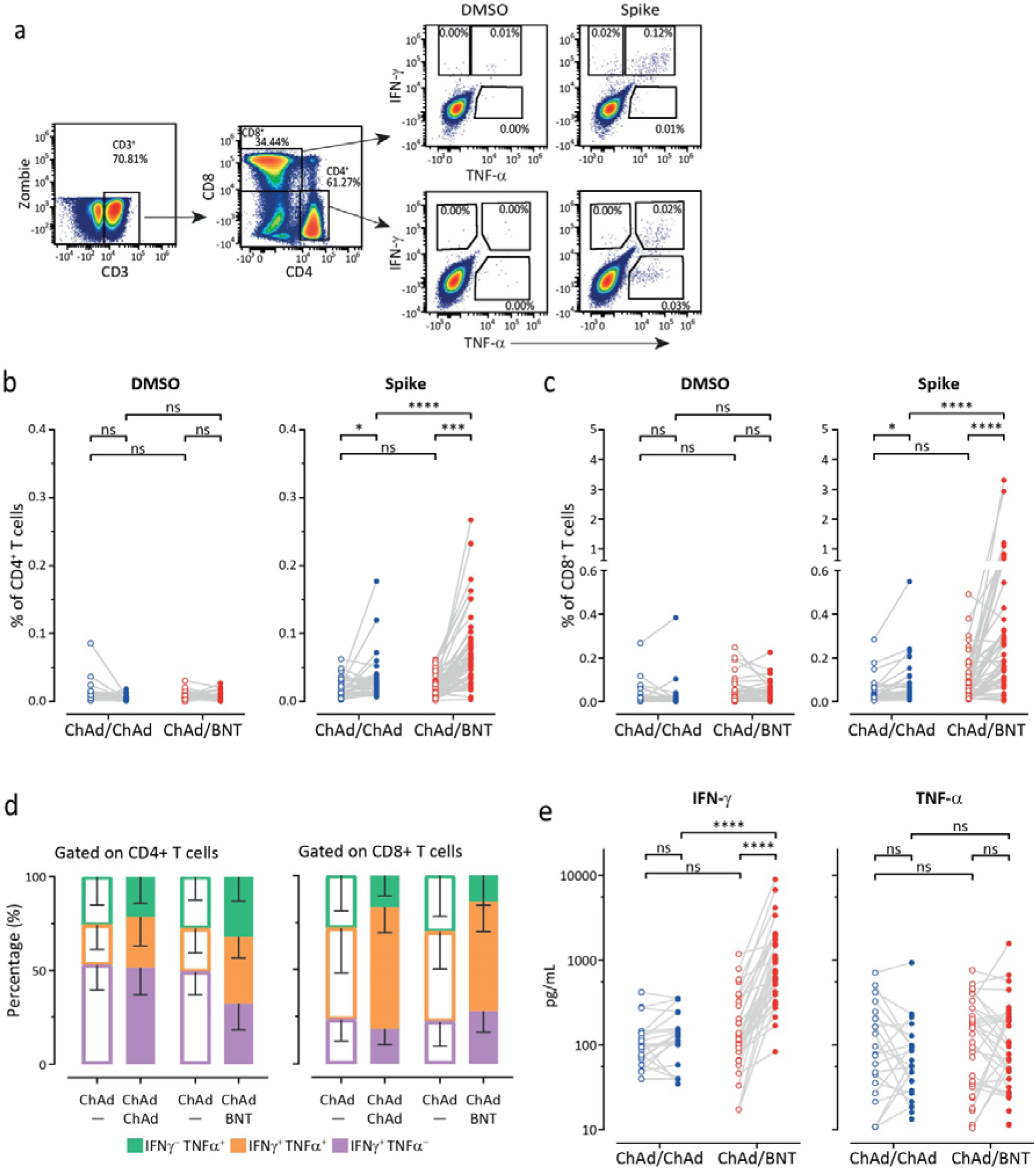
Heterologous ChAd / BNT vaccination induces stronger anti-SARS-CoV2 S T cell response than homologous ChAd / ChAd vaccination. a. Gating strategy used for detection of cytokine producing CD4^+^ and CD8^+^ T cells after *ex vivo* re-stimulation with DMSO or the pool of spike-specific peptides for 16hr. b. – c. Boost vaccination increased total percentage of cytokine-secreting CD4^+^ (b) and CD8^+^ (c) T cells. We calculated the total number of cytokine secreting cells as sum of IFN-γ^+^TNF-α^-^, IFN-γ^+^TNF-α^+^, IFN-γ^-^TNF-α^+^ cells in the gates indicated in a. d. Increased percentage of double-cytokine secreting CD4^+^ and CD8^+^ T cells after the second vaccine dose. e. IFN-γ and TNF-α concentration in full blood supernatants after stimulation with SARS-CoV-2 S1 domain for 20-24 h measured by LEGENDplex™ (Biolegend). Statistics: b., c., and e. **** p<0.001 Paired t test (within groups) or 2-way ANOVA followed by Sidak’s multiple comparison test (between groups).

The frequencies of S-specific CD4 T cells in blood samples collected before booster vaccination were significantly higher for both vaccination groups as compared to the MNE (control) peptides or DMSO alone (Fig. 2B, Extended Data Fig. 6). No significant differences were found between the ChAd/ChAd and the ChAd/BNT group (Fig. 2B, open circles). After boosting, the frequencies for S-specific CD4 T cells increased in both groups and were significantly higher in the ChAd/BNT group (Fig. 2B, filled dots). The same effect was observed for S-specific CD8 T cells. These cells were present at comparable frequencies in both groups prior boosting and increased in frequencies after boosting. Again, boosting with BNT induced higher frequencies than boosting with ChAd (Fig. 2C, filled dots). Regarding the distribution of S-specific CD8 T cells producing IFN-γ or TNF-α application of both booster vaccines led to an increase in the proportion of cells producing both cytokines simultaneously (Fig. 2D). Significant increase in S-specific IFN-γ release in the ChAd/BNT but not in the ChAd/ChAd group was confirmed independently (Fig. 2E).

Due to the abrupt recommendation of several European governments to discontinue the use of ChAd in the young- and middle-aged population, a unique situation was created in which heterologous prime-boost vaccination regimens were applied despite the lack of any information available regarding immunogenicity and safety aspects. Applying a broad array of tests, this study qualitatively and quantitatively assessed B- and T-cell mediated immune responses. It provides first insights into the immunogenic outcome of homologous and heterologous vaccination protocols with two vaccines – BNT and ChAd. Head-to-head comparison of ChAd-prime vaccinees who received either a ChAd or BNT booster immunization revealed that both regiments elicited additional immunity. Although this setup did not allow for randomization of the participants, our study unequivocally revealed that the group boosted with BNT showed stronger immune responses than the group boosted with ChAd. CD4 and CD8 T cell responses directed against S-protein epitopes were higher in frequencies and cells produced more IFN-γ upon re-stimulation. Likewise, the group boosted with BNT developed higher titers of anti-S-protein antibodies of both the IgG and IgA subclasses. It should be noted that these antibodies were highly efficient in neutralizing all three VoC tested in the present study. It had been reported before that vaccinees immunized with BNT/BNT also develop neutralizing antibodies against the VoC ^20^. Based on the data of n=46 participants of the CoCo Study cohort that were also immunized with BNT/BNT, we could confirm these findings in the present study. Our data also indicate that BNT/BNT and ChAd/BNT vaccinated individuals develop neutralizing antibodies to similar degrees two to three weeks after booster vaccination. Likewise, immune responses of the ChAd/ChAd group were in the range of earlier reported results ^11–13,21^. Although it would have been interesting to also characterize immune responses in a cohort of people immunized with BNT/ChAd, such individuals had not been available to us.

Extended studies, ideally including clinical endpoints, are needed to further characterize immune responses not only in heterologous immunized cohorts. It would be of particular importance to address for how long protective immune responses are maintained, first of all in people that are at elevated risk for developing severe COVID-19 but also in individuals that are known for mounting impaired immune responses.

## Data Availability

The data that support the findings of this study are available from the corresponding author upon reasonable request.

## Acknowledgements

This work was supported by the German Center for Infection Research TTU 01.938 (grant no 80018019238 to G.M.N.B and R.F) by Deutsche Forschungsgemeinschaft, (DFG, German Research Foundation) Excellence Strategy EXC 2155 “RESIST” to RF (Project ID39087428), by funds of the State of Lower Saxony (14-76103-184 CORONA-11/20) to RF, by funds of the BMBF (NaFoUniMedCovid19” FKZ: 01KX2021; Projects B-FAST) to RF and Deutsche Forschungsgemeinschaft, Project 158989968 - SFB 900/3 (Projects B1 to RF). We thank the CoCo Study participants for their support and the entire CoCo study team for help. We would like to thank Luis Manthey, Hannah Bartmann, Janine Topal, Kerstin Sträche, Birgit Heinisch, Michael Stephan, Mariel Nöhre, Simone Müller, Olivera Dragicevic, Anh Thu Tran, Kim Do Thi Hoang, Anna-Lena Boeck, Amy Kemp, and Inga Nehlmeier for technical and logistical support.

## Contributions

Study design: G.M.N.B and R.F.

Data collection: J.B.-M., S.I.H., A.C., I.O., M.V.S., G.M.R., A.D.-J., A.H., C.R., M.F., C. S.-F., I.R., S.W., A.B., J.M., J.R., A.J., G.S., G.B., J.M., M.H., S.P., T.K.

Data analysis: J.B.-M., S.I.H., A.C., I.O., M.H., B.B, M.V.S.

Data interpretation: B.B., R.F., G.M.N.B.

Writing: B.B., G.M.N.B., R.F. with comments from all authors.

