## Extended Data for "Humoral and cellular immune response against SARS-CoV-2 variants following heterologous and homologous ChAdOx1 nCoV-19/BNT162b2 vaccination"

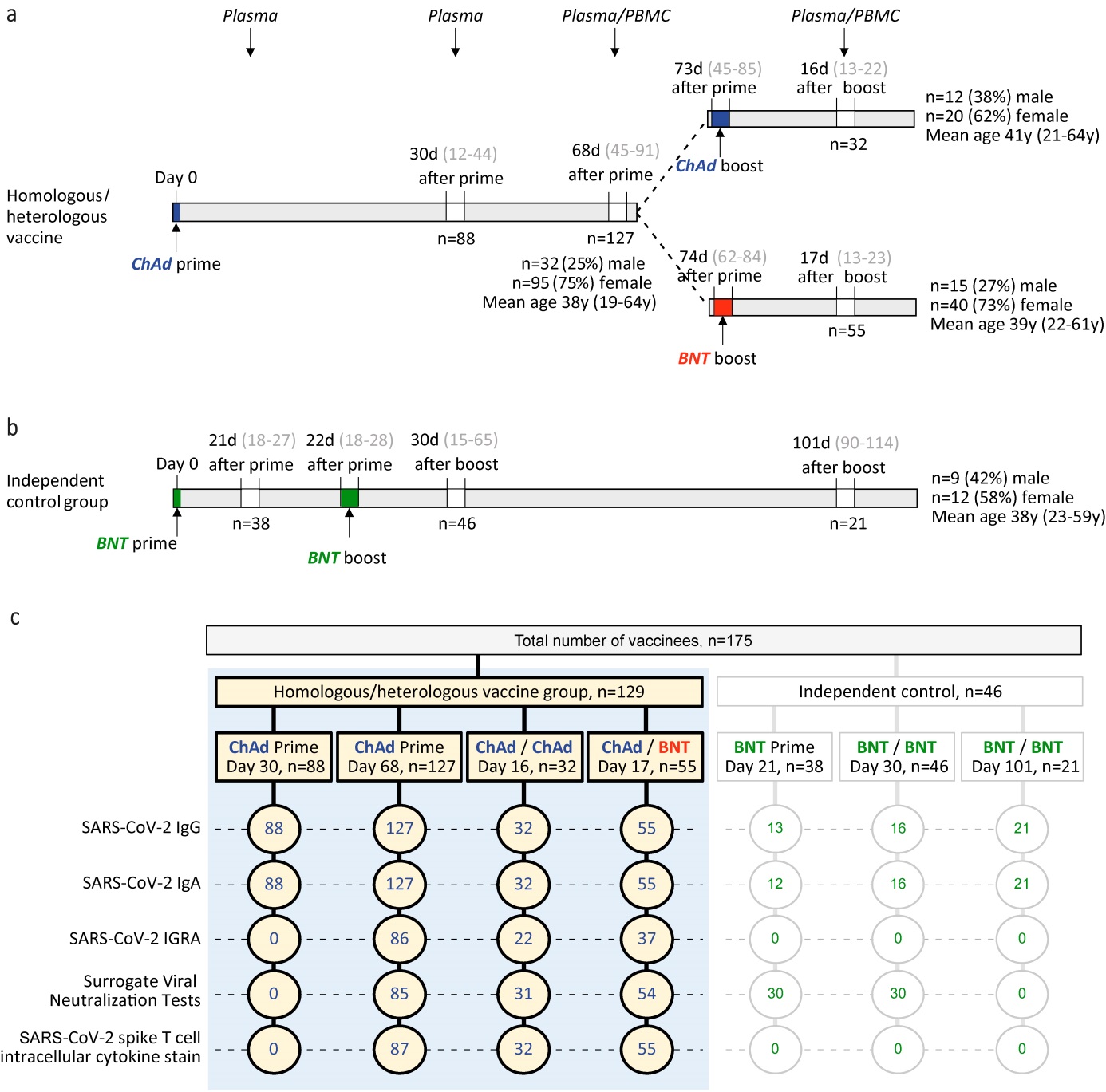

**Extended Data Figure 1.**

a. Participant recruitment scheme including age and sex information.
b. Participant recruitment scheme for vaccinees immunized twice with BNT vaccine.
c. Scheme indicating number of samples analyzed with each assay in each group.

**Extended Data Table 1.**

a. Time course of anti-S IgG and IgA decline over mean 38 days after ChAd prime and before full BNT vaccination.

| **ChAd prime** | Mean 30 d  after prime | Mean 68 d  after prime | Mean  reduction | P-value |
| --- | --- | --- | --- | --- |
| Anti-S IgG [RU/mL] ± SD | 92.7 ± 111.6 | 53.4 ± 53.5 | 43% | < 0.001 |
| Anti-S IgA ratio ± SD | 1.50 ± 1.74 | 0.52 ± .64 | 65% | < 0.001 |

b. Time course of anti-S IgG and IgA decline over mean 70 days in the independent control group after BNT/BNT full vaccination.

| **BNT/BNT** | Mean 31 d  after boost | Mean 101 d  after boost | Mean reduction | P-value |
| --- | --- | --- | --- | --- |
| Anti-S IgG [RU/mL] ± SD | 574.1 ± 199.9 | 303.2 ± 158.8 | 53% | < 0.001 |
| Anti-S IgA ratio ± SD | 5.06 ± 2.77 | 2.56 ± 2.29 | 49% | 0.005 |

c. Fold increase of anti-S IgG and IgA after ChAd prime (top values) followed by either ChAd or BNT booster vaccination (blue or red values).

| **ChAd** or **BNT boost** | Mean 18 d  after **ChAd** boost | Fold increase | Mean 16 d  after **BNT** boost | Fold increase |
| --- | --- | --- | --- | --- |
| Anti-S IgG [RU/mL] ± SD | 56.46 ± 104.73  160.9 ± 108.3 | 2.9 | 54.42 ± 48.63  625.7 ± 104.0 | 11.5 |
| Anti-S IgA ratio ± SD | 0.41±0.32  0.56 ± 0.59 | 2.2 | 0.56 ± 0.59  3.76 ± 2.23 | 6.7 |

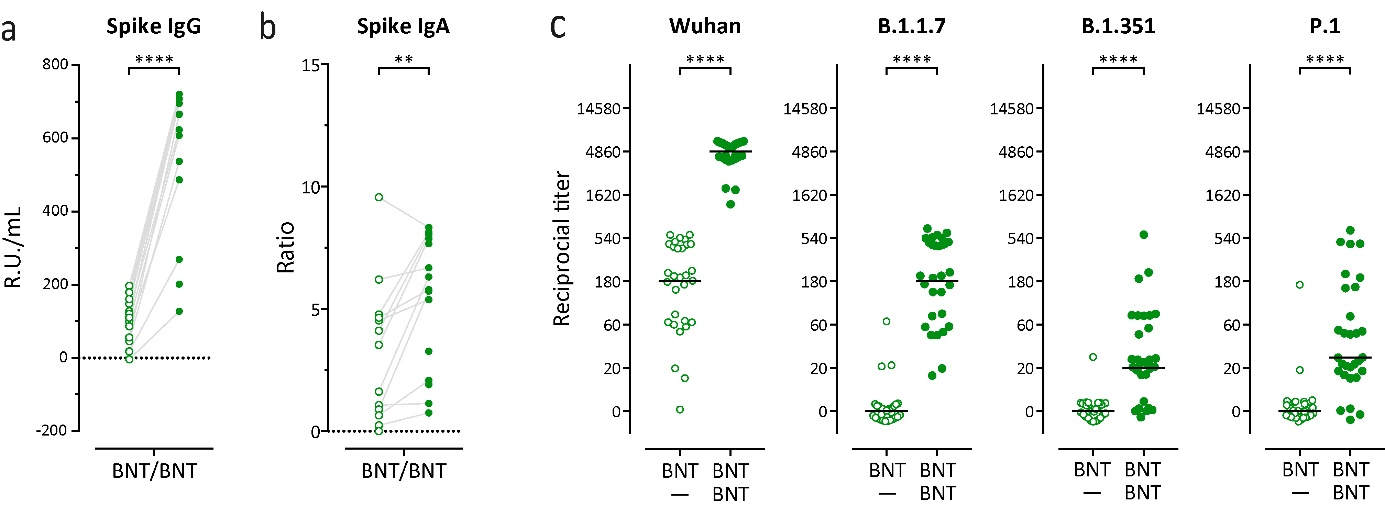

**Extended Data Fig. 2.** Humoral immune response against all SARS-CoV-2 variants following homologous BNT162b2 (BNT) / BNT162b2 (BNT) vaccination.

a. Spike-specific IgG and IgA levels in plasma after prime (open circles) and after booster (closed circles) homologous BNT/BNT vaccination.
b. Reciprocal titers of neutralizing antibodies against Wuhan, B.1.1.7-Spike (British), P.1-Spike (B.1.1.28.1; Brazilian), and B.1.351-Spike (South African) SARS-CoV2 variants measured using surrogate virus neutralization test (sVNT).

Statistics: a. **** p<0.001 Paired t test (within groups); b. **** p<0.001 Chi-square test for trend.

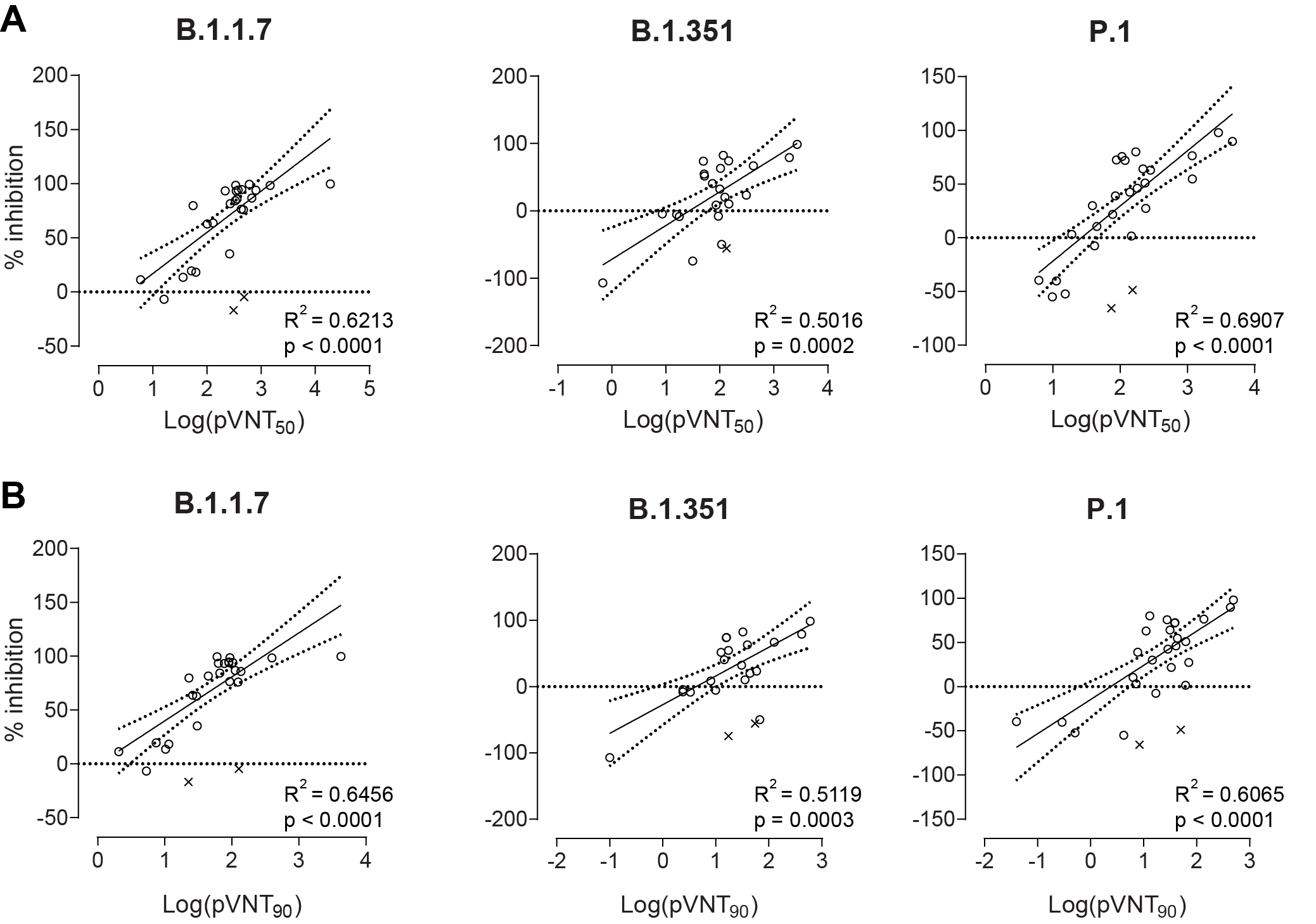

**Extended Data Fig. 3.** Efficacy of antibody neutralization against different SARS-CoV2 variants measured by surrogate virus neutralization test (sVNT) strongly positively correlates with the neutralization measured with pseudotyped virus neutralization test (pVNT).

Correlation (solid line) and 95% confidence intervals (dotted lines) between sVNT_1:20_ and antibody titers resulting in 50% (A) or 90% (B) reduction of luciferase in pVNT, indicated as pVNT_50_ and pVNT_90_, respectively. Dots, values from individual donors, outliers are marked with x and were defined as values with absolute residual value > 2SD of all residual values in each group of samples. Correlation was calculated using single linear regression.

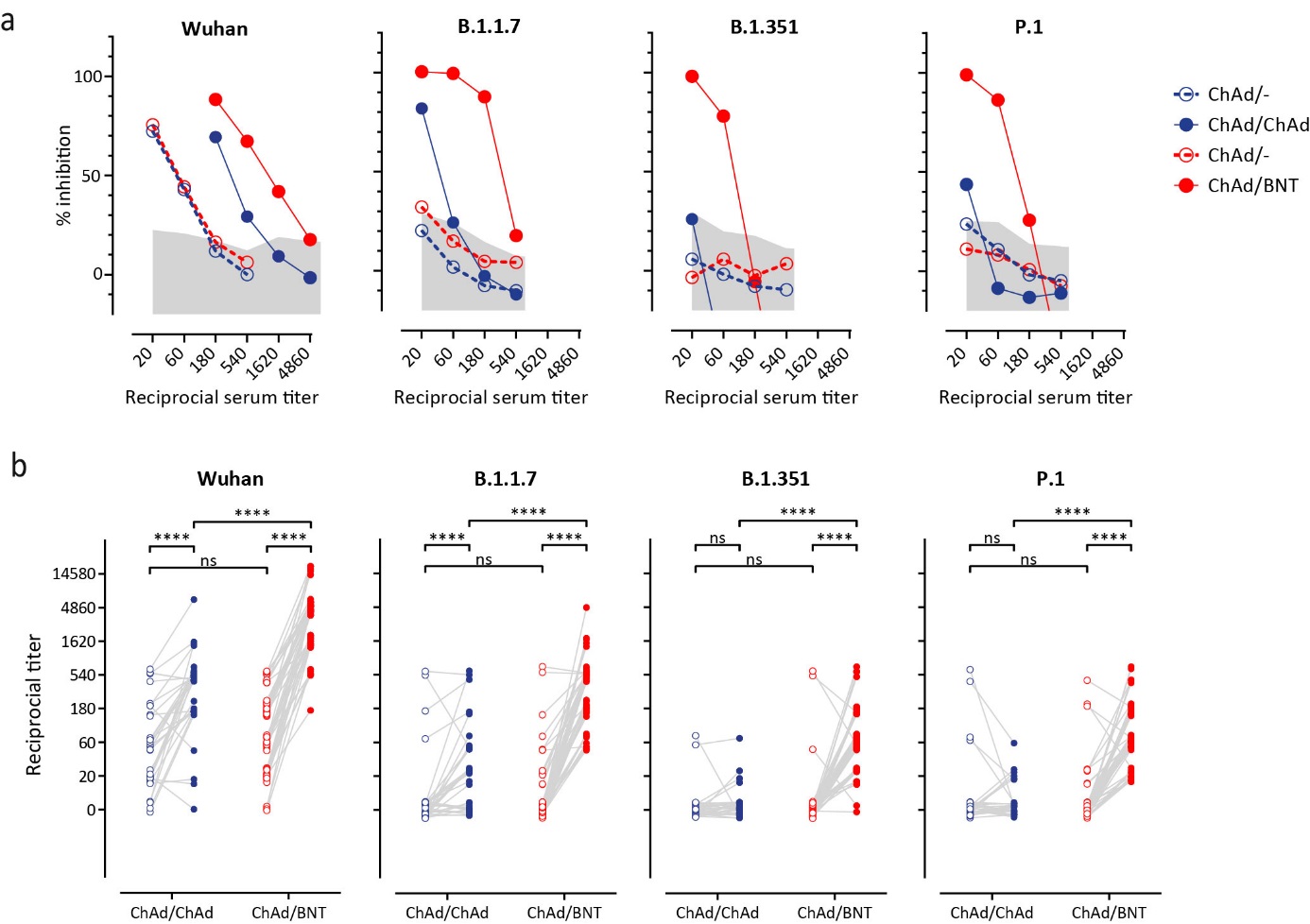

**Extended Data Fig. 4.** Surrogate virus neutralization test (sVNT) detects neutralizing antibodies interfering with binding of divergent SARS-CoV-2-S-RBD variants to human ACE2.

a. Inhibition of interaction of indicated SARS-CoV-2-S-RBD variants with ACE2 by the addition of plasma of vaccinee before (open circles) and after (closed circles) homologous ChAd/ChAd (blue symbols) and heterolgous ChAd/BNT (red symbols). Assay was performed in duplicates and shown as mean percentages of neutralization. Shaded areas represent mean + 2 SD of values from pre-COVID-19 plasma.
b. Reciprocal titers of neutralizing antibodies against Wuhan, B.1.1.7-Spike (British), P.1-Spike (B.1.1.28.1; Brazilian), and B.1.351-Spike (South African) SARS-CoV2 variants in plasma of each vaccinee before and after the boost immunization. **** p<0.001 Chi-square test for trend.

**Extended data Table 2.** List of reagents used for S-specific B cells flow cytometry.

| **Antigen** | **Conjugate** | **Clone** | **Order no.** | **Company** |
| --- | --- | --- | --- | --- |
| CD10 | PE | HI10a | 312204 | Biolegend |
| CD138 | PE-eF610 | MI15 | 61-1388-42 | Invitrogen |
| CD14 | BB700 | MφP9 | 566465 | BD |
| CD16 | BUV496 | 3G8 | 612944 | BD |
| CD19 | PerCP | HIB19 | 302228 | Biolegend |
| CD20 | BV421 | 2H7 | 302330 | Biolegend |
| CD21 | BUV661 | 1048 | 750187 | BD |
| CD24 | PE/Dazzle 594 | ML5 | 311134 | Biolegend |
| CD27 | BUV805 | L128 | 748704 | BD |
| CD3 | AF532 | UCHT1 | 58-0038-42 | Invitrogen |
| CD38 | PerCP-eF710 | HB7 | 46-0388-42 | Invitrogen |
| CD95 | PE-Cy5 | DX2 | 305610 | Biolegend |
| CXCR5 | BV750 | RF8B2 | 747111 | BD |
| IgD | BV480 | IA6-2 | 566138 | BD |
| IgM | Af647 | MHM-88 | 314536 | Biolegend |
| Viability | Zombie NIR^TM^ | - | 423106 | Biolegend |
| Anti-S BCR | mNeonGreen | Produced by T. Krey | | |

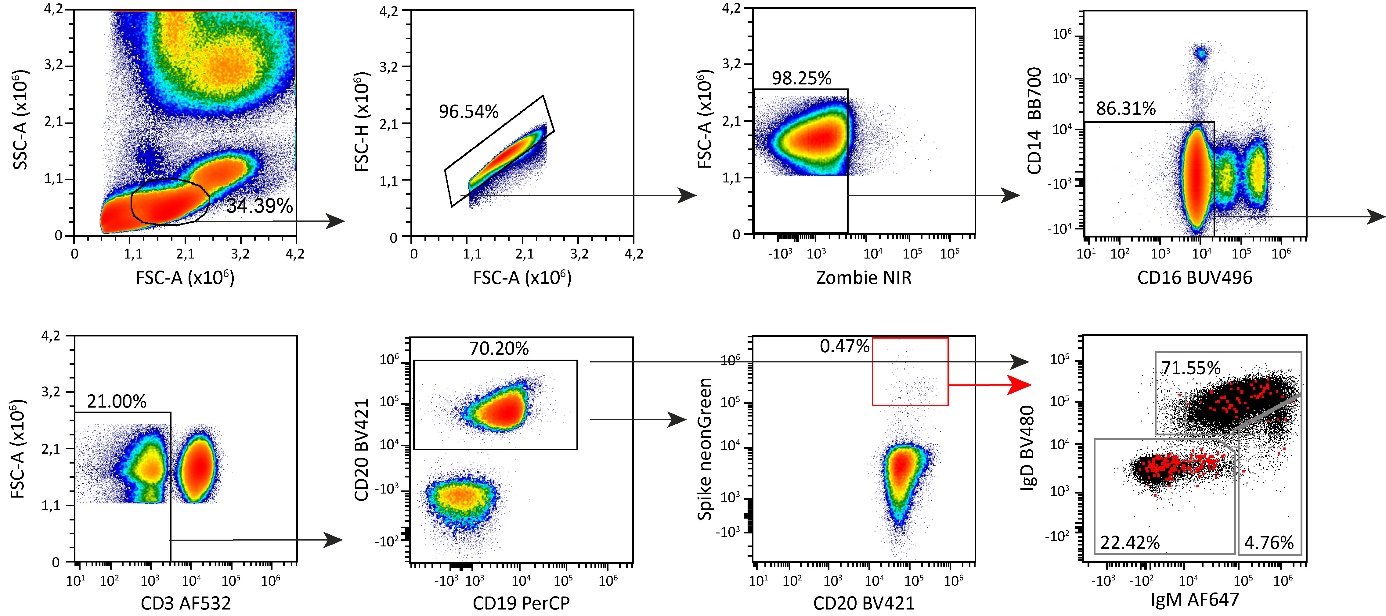

**Extended Data Fig. 5.** Gating strategy for SARS-CoV2-S (Spike)-specific B cell populations in blood using antibody panels from Extended Data Table 2. Pseudocolor plots show representative data from a female donor 71 days after priming with ChAd and 20 days post boost with BNT.

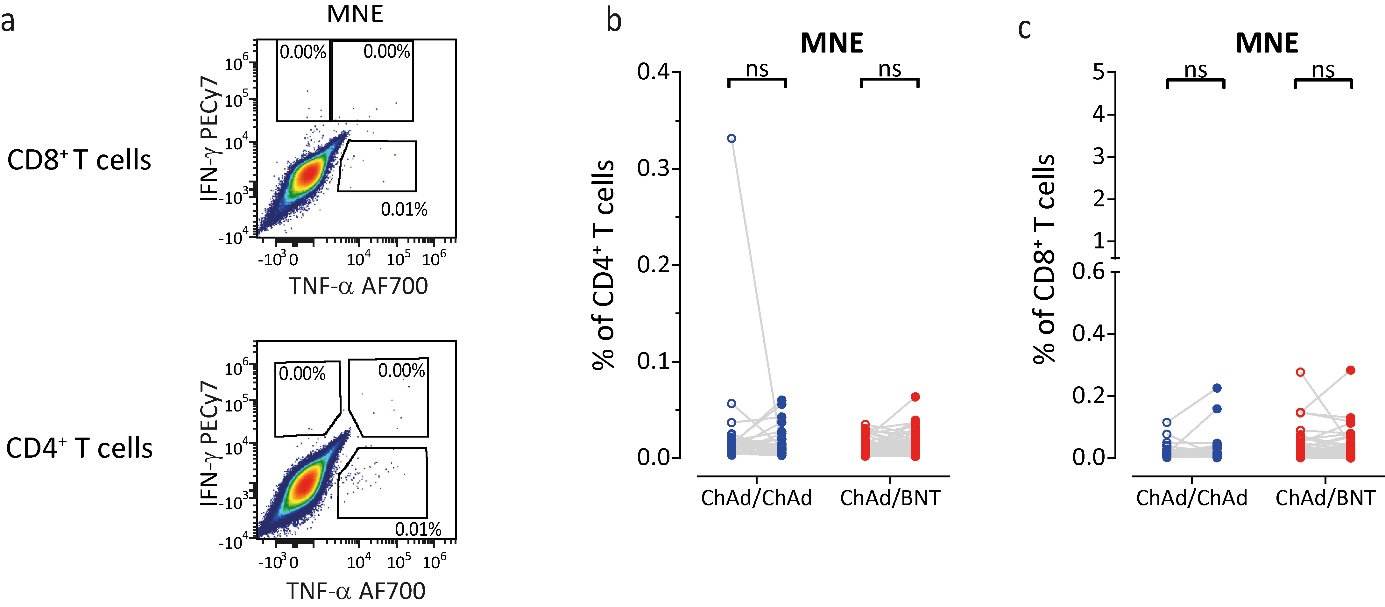

**Extended Data Fig. 6.** Cytokine production in T cells after 16 hr *ex vivo* restimulation with mixture of peptides from membrane (M), nucleocapsid (N), and envelope (E) SARS-CoV2 proteins. The total number of cytokine secreting cells calculated as sum of IFN-γ^+^TNF-α^-^, IFN-γ^+^TNF-α^+^, and IFN-γ^-^TNF-α^+^ cells as gated in (a) in CD4^+^ (b) and CD8^+^ (c) T cells. **** p<0.001 Paired t test (within groups).
