## Supplementary material for "Humoral and cellular immune response against SARS-CoV-2 variants following heterologous and homologous ChAdOx1 nCoV-19/BNT162b2 vaccination": Materials and Methods

**Online methods**

*Participants*

Participants for this analysis were from the COVID-19 Contact (CoCo) Study, which started in March 2020 and is an ongoing, prospective study monitoring anti-SARS-CoV-2 IgG immunoglobulin and immune responses in n=1493 health care professionals (HCP) at Hannover Medical School and individuals with potential contact to SARS-CoV-2^1^. An amendment from Dec 2020 allowed us to study the immune responses after COVID-19 vaccination. According to German regulations, HCP were prioritized for SARS-CoV2 vaccination and HCP at Hannover Medical School received first doses of the BNT or ChAd vaccine from Jan 6^th^, respectively Feb 16^th^, 2021 onwards. In general, booster vaccination took place about 21 days after BNT prime and around 2-3 months after ChAd prime. Booster vaccination of ChAd primed HCP started on May 3^rd^ and individuals could choose to receive either ChAd or BNT for second vaccination. We assumed that about 25% of all ChAd primed vaccinees would opt for a homologous booster. We estimated that a sample size of 30 subjects in each arm is sufficient to detect a clinically meaningful difference within each group assuming that specific IgG doubles from first vaccination (mean 95 RU/mL with a standard deviation of 113 RU/mL) using a two-tailed paired t-test of differences between means with 95% power and a 1% level of significance. The power calculation was performed with G*Power. Following written informed consent, we obtained peripheral blood samples by venipuncture. Individuals with previously PCR confirmed SARS-CoV-2 infection or SARS-CoV-2 seroconversion as determined by positive anti-SARS-CoV-2 NCP IgG before vaccinations were excluded from this analysis. Participants were 25% male and 75% female with a mean age of 38 years (range 19-64 years). After blood collection, we separated plasma from EDTA or lithium heparin blood (S-Monovette, Sarstedt) and stored it at minus 80°C until use. We used full blood or isolated PBMCs from whole blood samples by Ficoll gradient centrifugation and for stimulation with SARS-CoV-2 peptide pools.

*Pseudotyped virus neutralization assay (pVNT)*

pVNTs were performed at the Infection Biology Unit of the German Primate Center in Göttingen as described recently ^2^. Briefly, the rhabdoviral pseudotyped particles were produced in 293T cells transfected to express the desired SARS-CoV-2-S variant inoculated with VSV*DG-FLuc, a replication-deficient VSV vector that encodes for enhanced green fluorescent protein and firefly luciferase (FLuc) instead of VSV-G protein (kindly provided by Gert Zimmer, Institute of Virology and Immunology, Mittelhäusern, Switzerland). Produced pseudoparticles were collected, cleared from cellular debris by centrifugation and stored at -80 °C until used. For neutralization experiments, equal volumes of pseudotyped particles and heat-inactivated (56 °C, 30 min) plasma samples serially diluted in culture medium were mixed and incubated for 30 min at 37 °C. Afterwards, the samples together with non-plasma-exposed pseudotyped particles were used for transduction experiments. The assay was performed in 96-well plates in which Vero cells were inoculated with the respective pseudoytped particles/plasma mixtures. The transduction efficacy was analyzed at 16-18 hr post inoculation by measuring FLuc activity in lysed cells (Cell culture lysis reagent, Promega) using a commercial substrate (Beetle-Juice, PJK) and a plate luminometer (Hidex Sense Plate Reader, Hidex).

*Serology*

We determined SARS-CoV-2 IgG serology by quantitative ELISA (anti-SARS-CoV-2 S1 spike protein domain/receptor binding domain IgG SARS-CoV-2-QuantiVac, Euroimmun, Lübeck, Germany) according to the manufacturer’s instructions (dilution 1:400 or 1:600). We provide antibody levels expressed as RU/mL as assessed from a calibration curve with values above 11 RU/mL defined as positive. We performed anti-SARS-CoV-2 S1 spike protein domain IgA or anti SARS-CoV-2 nucleocapsid (NCP) IgG measurements according to the manufacturer’s instructions (Euroimmun, Lübeck, Germany) and expressed antibody amounts as IgA ratio (optical density divided by calibrator).

*Surrogate virus neutralization assay (sVNT) for SARS-CoV-2 variants*

To determine neutralizing antibodies against Wuhan-Spike, B.1.1.7-Spike (British), P.1-Spike (formerly named B.1.1.28.1; Brazilian), and B.1.351-Spike (South African) variants of SARS-CoV-2-S in plasma, we modified our recently established surrogate virus neutralization test (sVNT) ^3^. In detail, MaxiSorp 96F plates (Nunc) were coated with recombinant soluble hACE2-Fc(IgG1) protein at 300 ng per well in 50 μl coating buffer (30 mM Na_2_CO_3_, 70 mM NaHCO_3_, pH 9.6) at 4 °C overnight. After blocking with hACE2-Fc(IgG1), plates were washed with phosphate-buffered saline, 0.05% Tween-20 (PBST) and blocked with BD OptEIA Assay Diluent for 1.5 h at 37 °C. In the meantime, plasma samples were serially diluted threefold starting at 1:20 and then pre-incubated for 1 h at 37 °C with 1.5 ng recombinant SARS-CoV-2 Spike RBD of either the Wuhan strain (Trenzyme), the B.1.1.7 variant (N501Y), the B.1.351 variant (K417N, E484K, N501Y) or the P.1 variant (K417T, E484K, N501Y) (the latter three from SinoBiological), all with a C-terminal His-Tag. BD OptEIA Assay Diluent was used for preparing plasma sample as well as RBD dilutions. After pre-incubation with SARS-CoV-2 Spike RBDs, plasma samples were given onto the hACE2-coated MaxiSorp ELISA plates for 1 h at 37 °C. SARS-CoV-2 Spike RBDs pre-incubated with buffer only served as negative controls for inhibition. Plates were washed three times with PBST and incubated with an HRP-conjugated anti-His-tag antibody (clone HIS 3D5, provided by Helmholtz Zentrum München) for 1 h at 37 °C. Unbound antibody was removed by six washes with PBST. A colorimetric signal was developed on the enzymatic reaction of HRP with the chromogenic substrate 3,3',5,5'-tetramethylbenzidine (BD OptEIA TMB Substrate Reagent Set). An equal volume of 0.2 M H_2_SO_4_ was added to stop the reaction, and the absorbance readings at 450 nm and 570 nm were acquired using a SpectraMax iD3 microplate reader (Molecular Devices). For each well, the percent inhibition was calculated from optical density (OD) values after subtraction of background values as: Inhibition (%) = (1 − Sample OD value/Average SARS-CoV-2 S RBD OD value) × 100. Neutralizing sVNT titers were determined as the dilution with binding reduction > mean + 2SD of values from a plasma pool consisting of three pre-pandemic plasma samples.

*SARS-CoV-2 protein peptide pools*

We ordered 15 aminoacid (aa) long and 10 aa overlapping peptide pools spanning the whole length of SARS-CoV2-S (total 253 peptides), -M (43 peptides), -N (82 peptides) or –E (12 peptides; peptide no 4 could not be synthesized) from GeneScript. All lyophilized peptides were synthesized at >95% purity and reconstituted at a stock concentration of 50 mg/mL in DMSO (Sigma-Aldrich), except for 9 SARS-CoV2-S overlapping peptides (number 24, 190, 191, 225, 226, 234, 244, 245 and 246), 2 for SARS-CoV2-M (number 15 and 16), 1 for SARS-CoV2-N (number 61) and all 12 SARS-CoV2-E peptides that were dissolved at 25 mg/mL due to solubility issues. All peptides in DMSO stocks were stored at -80°C until used.

*T cell re-stimulation assay*

PBMCs, isolated using a Ficoll gradient, were re-suspended at concentration of 20 x 10^6^ cells/ml in complete RPMI medium [RPMI 1640 (Gibco) supplemented with 10% FBS (GE Healthcare Life Sciences, Logan, UT), 1mM sodium pyruvate, 50 µM β-mercaptoethanol, 1% streptomycin/penicillin (all Gibco)]. For stimulation, cells were diluted with equal volume of peptide pools containing S-protein or mixture of M-, N- and E-proteins. Peptide pools were prepared in complete RPMI containing brefeldin A (Sigma-Aldrich) at final concentration of 10 µg/ml. In the final mixture each peptide had concentration of 2 µg (~1.2 nmol)/ml, except for SARS-CoV2-S peptides number 24, 190, 191, 225, 226, 234, 244, 245 and 246, SARS-CoV2-M peptides 15 and 16, and SARS-CoV2-N peptide 61, which were used at final concentration of 1 µg/ml due to solubility issues. As a negative control, we stimulated the cells with DMSO, used in maximal volume corresponding to DMSO amount in peptide pools (5 % v/v). In each experiment, we used cells stimulated with Phorbol-12-myristate-13-acetate (PMA; Calbiochem) and ionomycin (Invitrogen) at final concentration of 50 ng/mL and 1500 ng/mL, respectively, as an internal positive control. Cells were then incubated for 12-16 hr at 37°C, 5% CO_2_. After washing, cells were resuspended in MACS buffer (PBS supplemented with 3% FBS and 2mM EDTA). Non-specific antibody binding was blocked by incubating samples with 10% mouse serum at 4°C for 15 minutes. Next, without washing, an antibody mix of anti-CD3-AF532 (UCHT1; #58-0038-42; Invitrogen), anti-CD4-BUV563 (SK3; #612913; BD Biosciences), anti-CD8-SparkBlue 550 (SK1; #344760; Biolegend), anti-CD45RA (HI100, #740298, BD Biosciences), anti-CCR7 (G043H7 , #353230, Biolegend), anti-CD38 (HB7, #46-0388-42, Invitrogen) and Zombie NIR™ Fixable Viability Kit (#423106, BioLegend) was added. After staining for 20 minutes at RT, cells were washed before they were fixed and permeabilized (#554714, BD Biosciences) according to the manufacturers’ protocol. Next, intracellular cytokines were stained using anti-IFN-PE-Cy7 (4S.B3, #502528, Biolegend), anti-TNF-AF700 (Mab11; #561023; BD Biosciences) and anti-IL-17A-BV421 (TC11-18H10.1; #506926, Biolegend) for 45 minutes on RT. Excess antibodies was washed away and cells were the acquired on Cytek Aurora spectral flow cytometer (Cytek) equipped with five lasers operating on 355nm, 405nm, 488nm, 561nm and 640nm. All flow cytometry data was analyzed using FCS Express V7 (Denovo).

*Flow cytometric analysis of S-specific B cells*

Total leukocytes were isolated from whole blood using erythrolysis in 0.83% ammonium chloride solution. Isolated cells were then washed, counted and resuspended in PBS and stained for 20 mins on RT with an antibody mix containing antibodies listed in Extended Data Table 2 together with Spike-mNEONGreen protein (5μg per reaction; production will be described elsewhere). After one wash, samples were acquired on spectral flow cytometer and the data was analyzed as described above.

*Quantification of IFNγ and TNFα release*

0.5 mL full blood were stimulated with manufacturer’s selected parts of the SARS-CoV-2 S1 domain of the Spike Protein for a period of 20-24 h. We carried out negative and positive controls according to the manufacturer’s instruction (SARS-CoV-2 Interferon Gamma Release Assay, IGRA (Euroimmun, Lübeck, Germany). Following stimulation, supernatants were collected after centrifugation and examined by LEGENDplex™ kit (BioLegend) according to the manufacturer’s instructions. Data were acquired with a LSR II flow cytometer (BD Biosciences) using BD’s FACSDiva v8.0.1 Software and analyzed with the LEGENDplex™ Data Analysis Software Suite.

**Ethics committee approval.** The CoCo Study (German Clinical Trial Register DRKS00021152) was approved by the Internal Review Board of Hannover Medical School (institutional review board no. 8973_BO-K_2020, amendment Dec 2020).
